# CASCADIA: A prospective community-based study protocol for assessing SARS-CoV-2 vaccine effectiveness in children and adults utilizing a remote nasal swab collection and web-based survey design

**DOI:** 10.1101/2023.01.05.22283913

**Authors:** Tara M. Babu, Leora R. Feldstein, Sharon Saydah, Zachary Acker, Cassandra L. Boisvert, Melissa Briggs-Hagen, Marco Carone, Amanda Casto, Sarah N. Cox, Brenna Ehmen, Janet A. Englund, Stephen P. Fortmann, Collrane J. Frivold, Holly Groom, Peter Han, Jennifer L. Kuntz, Tina Lockwood, Claire M. Midgley, Richard A. Mularski, Tara Ogilvie, Sacha Reich, Mark A. Schmidt, Ning Smith, Lea Starita, Jeremy Stone, Meredith Vandermeer, Ana A. Weil, Caitlin R. Wolf, Helen Y. Chu, Allison L. Naleway

**Author notes:** Corresponding Author: Tara Babu, University of Washington. This activity was reviewed by CDC and was conducted consistent with applicable federal law and CDC policy. The findings and conclusions in this report are those of the authors and do not necessarily represent the views of the Centers for Disease Control and Prevention.

## Abstract

**Introduction:** Although SARS-CoV-2 vaccines were first approved under Emergency Use Authorization by the FDA in late 2020 for adults, approval for young children 6 months to < 5 years of age did not occur until 2022. Understanding real world vaccine effectiveness in the setting of emerging variants is critical. The primary goal of this study is to evaluate SARS-CoV-2 vaccine effectiveness (VE) against infection among children aged >6 months and adults aged <50 years.

**Methods:** CASCADIA is a four-year community-based prospective study of SARS-CoV-2 VE among adult and pediatric populations aged 6 months to 49 years in Oregon and Washington. At enrollment and regular intervals, participants complete a sociodemographic questionnaire. Individuals provide a blood sample at enrollment and annually thereafter, with additional, optional blood draws after infection and vaccination. Participants complete weekly self-collection of anterior nasal swabs and symptom questionnaires. Swabs are tested for SARS-CoV-2 and other respiratory pathogens by RT-PCR, with results of selected pathogens returned to participants; nasal swabs with SARS-CoV-2 detected will undergo whole genome sequencing. Participants who report symptoms outside of their weekly swab collection and symptom survey are asked to collect an additional swab. Participants who test positive for SARS-CoV-2 undergo serial swab collection every three days for three weeks. Serum samples are tested for SARS-CoV-2 antibody by binding and neutralization assays.

**Analysis:** Cox regression models will be used to estimate the hazard ratio associated with SARS-CoV-2 vaccination among the pediatric and adult population, controlling for demographic factors and potential confounders, including clustering within households.

**Ethics and dissemination:** All study materials including the protocol, consent forms, participant communication and recruitment materials, and data collection instruments were approved by the Kaiser Permanente Northwest (KPNW) Institutional Review Board, the IRB of record for the study.

**Strengths/Limitations:**

- CASCADIA will include a large sample of children and adults that will contribute to estimation of vaccine effectiveness.
- The study will generate a data repository that can be used to address many research questions, such as duration of SARS-CoV-2 serologic results, post-acute sequelae of COVID-19, and re-infection rates.
- Retention and compliance may be challenging given the four-year duration of the study.
- Annual blood collection for assessment of humoral immunity may be a potential deterrent for participation, particularly among younger children.

## INTRODUCTION

Severe acute respiratory syndrome coronavirus 2 (SARS-CoV-2), the virus that causes coronavirus disease 2019 (COVID-19) rapidly disseminated across the world and was declared a pandemic by the World Health Organization on March 11, 2020.^1^ As of November 2022, COVID-19 has resulted in over 631, 000, 000 cases and 6,500,000 deaths worldwide.^2^ Currently, in the United States over 97,000,000 cases of COVID-19 have been reported, resulting in over 1 million deaths.^2^

As of October 27, 2022,, approximately 14.8 million children in the United States have tested positive for SARS-CoV-2 as reported to state health departments, accounting for 18.2% of all COVID-19 cases in those states.^3^ Throughout the pandemic, pediatric infections have been milder than infections among adults, and have resulted in a lower frequency of hospitalizations, deaths, and post-COVID conditions.^4 5 6^ However, disease burden has varied by variant strain. For instance, the emergence of the B.1.1.529 (Omicron) variant in the US from September 2021-January 2022 resulted in a 2.3 times higher peak hospitalization rate among children aged 5-11 years than the peak rate during the B.1.617.2 (Delta) variant surge.^7^ With emerging variants of concern (VOCs), understanding infection and reinfection rates, and variant specific outcomes, will be important for future prevention planning.^8-15^

During the COVID-19 pandemic, a worldwide collaborative effort resulted in the development of COVID-19 vaccines shown to decrease mortality and severe morbidity from SARS-CoV-2 infection.^16 17 18 19^ Due to emerging VOCs, waning antibody levels, and increased neutralizing antibody response following booster vaccines, booster vaccines have been implemented as a strategy for prevention of COVID-19 disease in several countries including the United States.^20 21^ VE data in the setting of novel VOCs may guide future vaccination strategies.

From November 2021 to June 2022, only the Pfizer-BioNTech vaccine had received authorization for use among children aged ≥5 years for a primary vaccine series.^22^ In May 2022, a booster vaccine was approved under EUA for 5 years and older. ^23^ On June 17, 2022, both BNT162b2 [Pfizer-BioNTech] and mRNA-1273 vaccine [Moderna] received approval under EUA for children 6 months and older.^24^ According to the CDC, 68% of the US population over the age of 5 years have received a COVID-19 primary vaccine series.^25^ However, rates of vaccine uptake vary by age strata. According to the CDC reported primary vaccine series completion stratified by age includes 1.7% in children <2 years, 3.2% in 2-4 years, 31.6% in 5-11 years, 60.8% in 12-17 years, 65.5% in 18-24 years, and 71% in 25-49 years. The rate of first booster vaccine uptake is reported as 15.6% in children 5-11 years, 29.3% in 12-17 years, 34.2% in 18-24 years, and 41.5% in 25-49 years.^25^ Many children remain unvaccinated. Additionally, parental/guardian vaccine hesitancy may delay pediatric vaccine uptake. As the landscape evolves with vaccine uptake, new emerging variants with potential for immune escape, and the introduction of novel vaccines, a longitudinal surveillance study will inform our understanding of vaccine effectiveness.

In the CASCADIA study, active surveillance and testing protocols will systematically identify both asymptomatic and symptomatic SARS-CoV-2 infection. Enrolling a cohort of children as soon as possible after the recent EUA of bivalent pediatric vaccines enables us to estimate the real-world COVID-19 VE in a pediatric population. The serology components of the protocol will allow for the evaluation of humoral immune responses to SARS-CoV-2 infection and vaccination and will allow for comparisons of symptomatic and asymptomatic infections and for primary infection vs. reinfection over time. The study data will aid in informing public health measures and vaccine strategies.

### AIMS

The primary study objective is to estimate the effectiveness of authorized COVID-19 vaccines against laboratory-confirmed symptomatic and asymptomatic SARS-CoV-2 virus infection among children aged 6 months to 17 years. A secondary objective is to estimate VE among adults 18-49 years of age. Additional primary and secondary objectives include evaluating humoral immune response following vaccination and infection, estimating secondary attack rates within households, and monitoring evolution of the SARS-CoV-2 virus by genomic sequencing (Table 1). Exploratory objectives include evaluating the incidence and outcomes of other respiratory viruses including respiratory syncytial virus (RSV) and influenza (Supplemental Table 1).

**Table 1.**
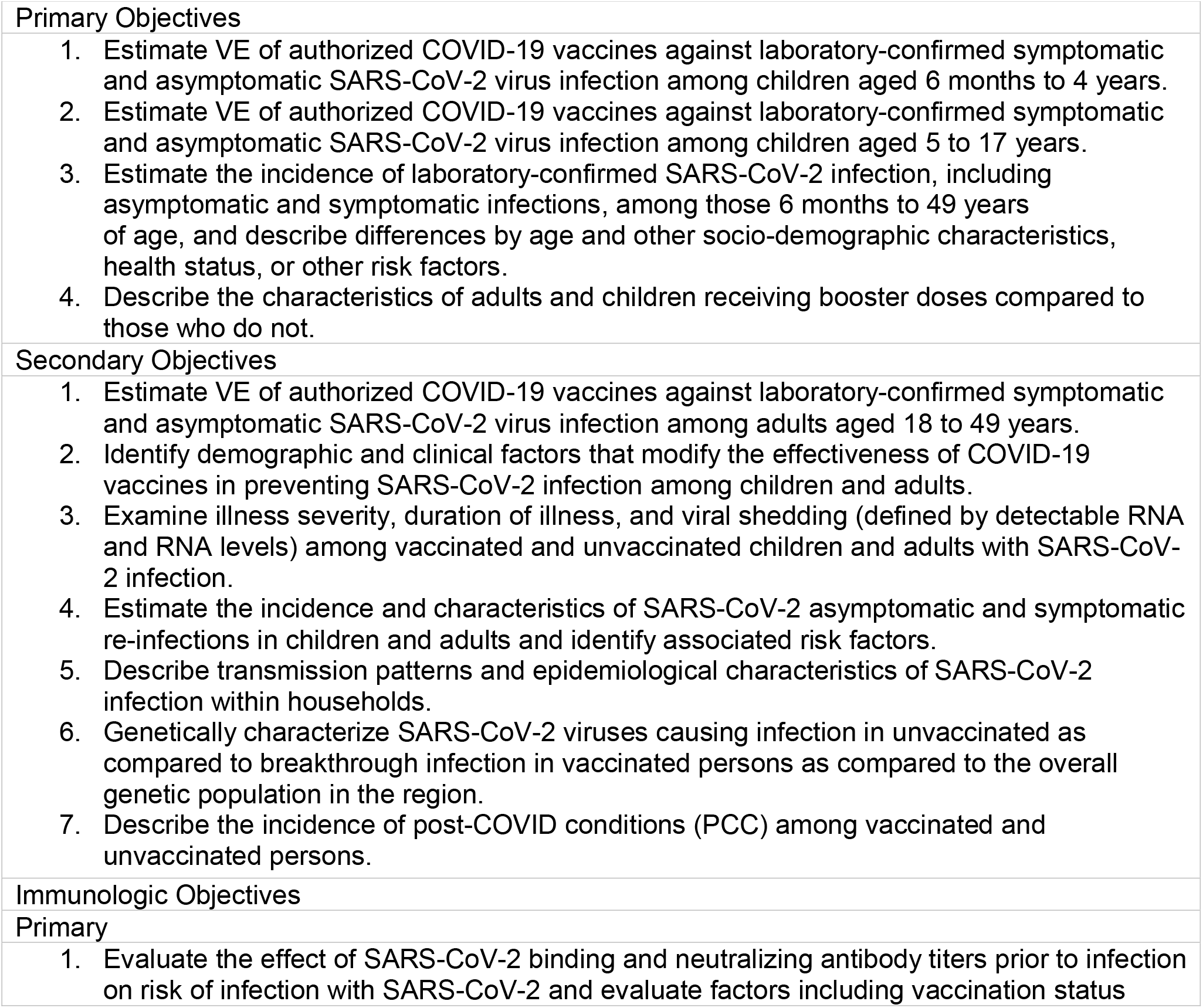
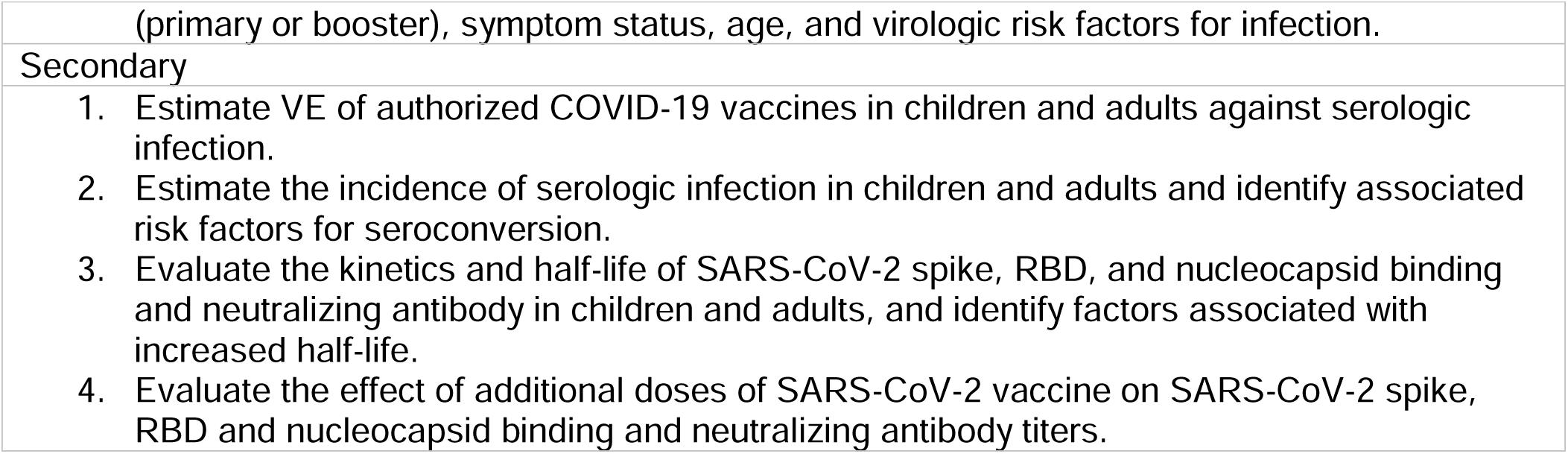
CASCADIA Study Objectives

| Table 1. CASCADIA Study Objectives |
| --- |
| Primary Objectives |
| 1. Estimate VE of authorized COVID-19 vaccines against laboratory-confirmed symptomatic and asymptomatic SARS-CoV-2 virus infection among children aged 6 months to 4 years.<br>2. Estimate VE of authorized COVID-19 vaccines against laboratory-confirmed symptomatic and asymptomatic SARS-CoV-2 virus infection among children aged 5 to 17 years.<br>3. Estimate the incidence of laboratory-confirmed SARS-CoV-2 infection, including asymptomatic and symptomatic infections, among those 6 months to 49 years of age, and describe differences by age and other socio-demographic characteristics, health status, or other risk factors.<br>4. Describe the characteristics of adults and children receiving booster doses compared to those who do not. |
| Secondary Objectives |
| 1. Estimate VE of authorized COVID-19 vaccines against laboratory-confirmed symptomatic and asymptomatic SARS-CoV-2 virus infection among adults aged 18 to 49 years.<br>2. Identify demographic and clinical factors that modify the effectiveness of COVID-19 vaccines in preventing SARS-CoV-2 infection among children and adults.<br>3. Examine illness severity, duration of illness, and viral shedding (defined by detectable RNA and RNA levels) among vaccinated and unvaccinated children and adults with SARS-CoV-2 infection.<br>4. Estimate the incidence and characteristics of SARS-CoV-2 asymptomatic and symptomatic re-infections in children and adults and identify associated risk factors.<br>5. Describe transmission patterns and epidemiological characteristics of SARS-CoV-2 infection within households.<br>6. Genetically characterize SARS-CoV-2 viruses causing infection in unvaccinated as compared to breakthrough infection in vaccinated persons as compared to the overall genetic population in the region.<br>7. Describe the incidence of post-COVID conditions (PCC) among vaccinated and unvaccinated persons. |
| Immunologic Objectives |
| Primary |
| 1. Evaluate the effect of SARS-CoV-2 binding and neutralizing antibody titers prior to infection on risk of infection with SARS-CoV-2 and evaluate factors including vaccination status |
| (primary or booster), symptom status, age, and virologic risk factors for infection. |
| Secondary |
| 1. Estimate VE of authorized COVID-19 vaccines in children and adults against serologic infection.<br>2. Estimate the incidence of serologic infection in children and adults and identify associated risk factors for seroconversion.<br>3. Evaluate the kinetics and half-life of SARS-CoV-2 spike, RBD, and nucleocapsid binding and neutralizing antibody in children and adults, and identify factors associated with increased half-life.<br>4. Evaluate the effect of additional doses of SARS-CoV-2 vaccine on SARS-CoV-2 spike, RBD and nucleocapsid binding and neutralizing antibody titers. |

## METHODS AND ANALYSIS

### Setting

The CASCADIA study is being conducted in Washington (WA) and Oregon (OR) through a collaboration between Kaiser Permanente Northwest (KPNW), University of Washington Medicine, and Seattle Children’s Hospital (SCH).

### Study Design

CASCADIA is a prospective longitudinal cohort study of children and adults designed to evaluate VE against laboratory-confirmed SARS-CoV-2 infection, and to estimate the incidence of asymptomatic and symptomatic SARS-CoV-2 infection. Approximately 2,400 children aged 6 months to 17 years and 1,100 adults aged 18 to 49 years will be enrolled and followed for up to four years; the study population will be split equally between KPNW and UW/SCH. Households with multiple participants residing together will be prioritized for enrollment. The CASCADIA study is funded by the US Centers for Disease Control and Prevention (CDC). Investigators from KPNW, UW, and SCH, in consultation with CDC, provided input on the study design, data collection instruments, laboratory testing procedures, data analysis, and implementation of this study. All sites utilize the same data collection instruments, and the protocol and study materials were reviewed and approved by the KPNW Institutional Review Board.

### Recruitment

In the preceding months to study launch, preparation included reinitiating and building new relationships with community partners. Prior to the official study launch, a soft launch was conducted targeting 3-5 families per site (KPNW and UW) and feedback was collected prior to beginning general study enrollment. The soft launch led to improvements in participant facing materials and contributed to the broader recruitment strategy (Figure 1). Recruitment strategies involve outreach to KPNW health plan members and local school districts and daycares; press releases and social media campaigns; and outreach to community-based organizations and other health care partners, prioritizing those with strong ties to racial and ethnic minorities and low-income individuals, where possible. Recruitment activities will continue during the study to replace participants who withdraw and adapted to meet demographic and vaccination enrollment targets.

**Figure 1.**
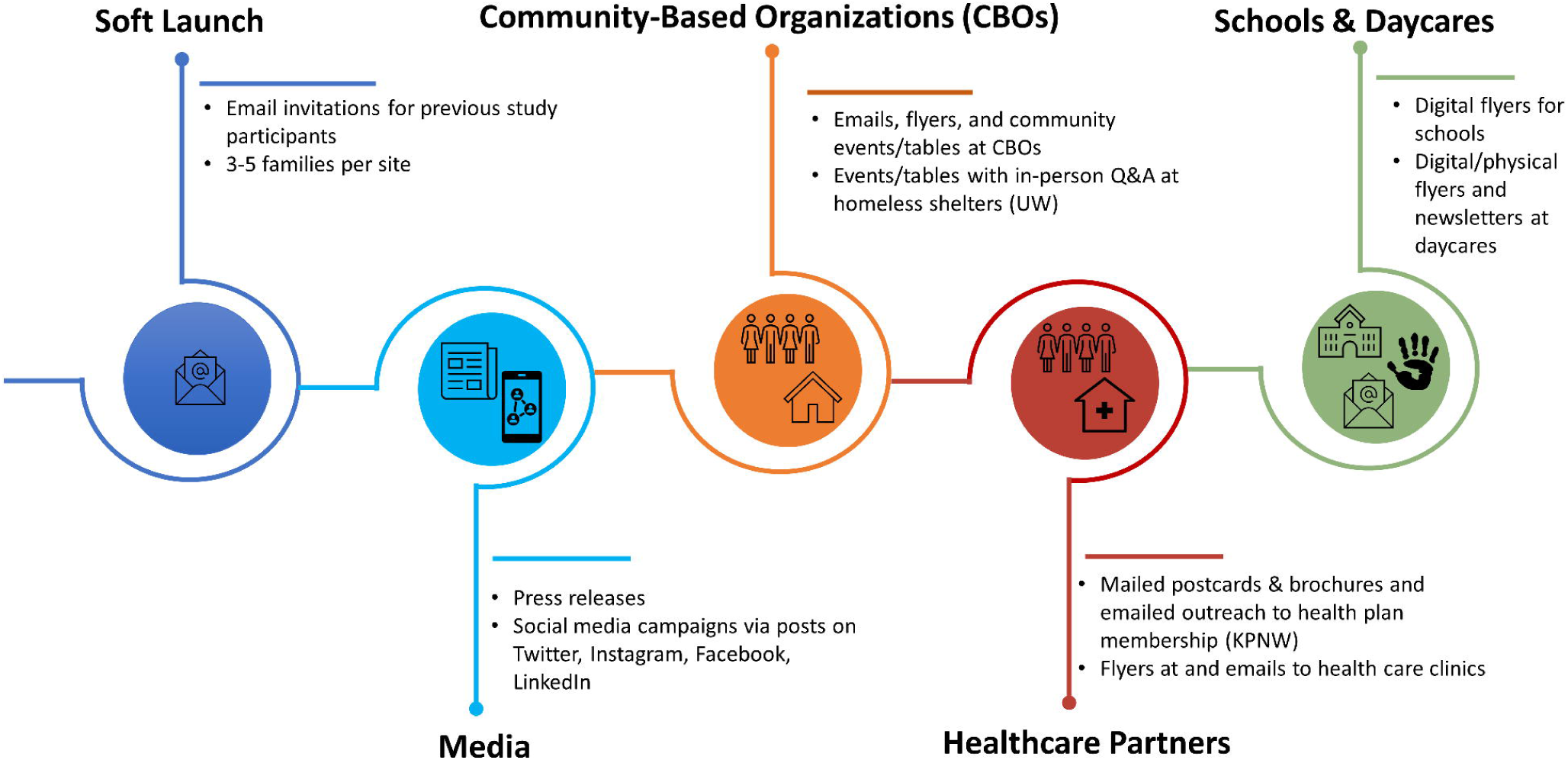
Tiered Recruitment Strategy

### Data Collection

Data collection and survey implementation is performed through Research Electronic Data Capture (REDCap), a metadata-driven EDC software and database methodology designed at Vanderbilt University.^26^ Web-based surveys are completed by participants and are entered directly into the study REDCap database.

### Screening

A central study website (www.cascadiastudy.org) enables potential participants to review information about the study and eligibility criteria (Table 2) and watch a video that provides an overview of the study and explains what is involved in participating in the study. The website links to an eligibility screener that collects names, age, sociodemographic data, contact information, enrollment site, and number of COVID-19 vaccines received. The website is available in English and will be available Spanish. Study staff monitor data from the eligibility screener and eligible participants are contacted and scheduled for consent/assent and enrollment visits. Eligible participants are offered the opportunity to self-schedule their consent call or enrollment visit. Once enrolled in the study, participants are granted access to a secure household home page through REDCap and are assigned a unique participant identification (PTID) number for the duration of the study.

**Table 2.** Eligibility Criteria

|  |
| --- |
| *Inclusion criteria* |
| Age ≥ 6 months to 49 years on the date of enrollment |
| Access to the internet and telephone |
| Willingness and ability to complete periodic data collection activities online |
| Willingness to collect □ nasal swabs □ (either self-collected □ or collected by □ the □ child's □ parent/guardian) □ and arrange pick-up time and location at least once per □ week for pick up and delivery of swabs |
| Willingness to participate in an enrollment blood collection and □ a subsequent minimum of one blood collection per year |
| Willing and able to provide consent or assent to participate |
| Live within the Kaiser Permanente Northwest catchment area of Oregon and southern Washington, or within the University of Washington catchment area of King, Snohomish, and Pierce Counties in Washington State |
| *Exclusion criteria* |
| Participation in a clinical trial of investigational prevention therapies for SARS- CoV-2 infections, such as prophylactic antiviral medications or other immune system modifying interventions in the 3 months prior to enrollment |
| Unwilling or unable to provide consent or assent to participate |
| Registered in Kaiser Permanente Northwest exclusion database |
| Unable to comply with study procedures, as determined by the investigators |

### Enrollment and Surveillance

Following the informed consent/assent process, participants are required to have or attempt a baseline blood draw and asked to complete an online enrollment survey (Supplemental Table 2). Web based surveys are utilized to collect data throughout the duration of the study with varying content by survey (Supplemental Table 3). Study procedures including surveys and swabbing are demonstrated in Figures 2A&2B (also see Supplemental Table 4).

**Figure 2.**
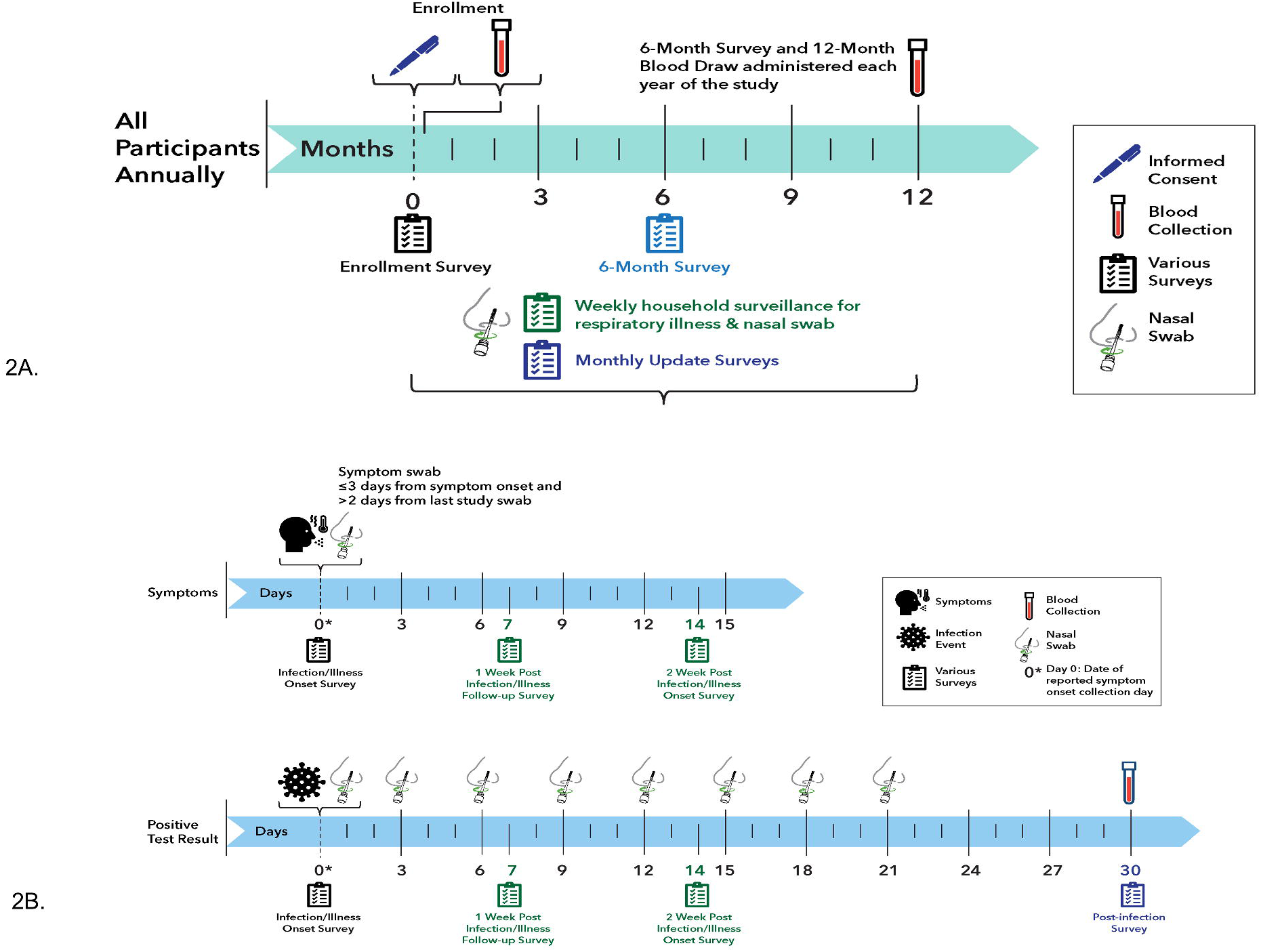
2A. CASCADIA Annual Study Procedures, 2B. Participant flow for COVID-19 like symptoms or positive SARS-CoV-2 testing

### Weekly Surveys

Participants or their parent/guardian complete a weekly Symptom & Swabbing survey which asks if they have experienced ≥ 1 of the following symptoms for over 24 hours: fever or chills, cough, shortness of breath or difficulty breathing, fatigue, muscle or body aches, headache, new loss of taste or smell, sore throat, congestion or runny nose, nausea or vomiting, diarrhea, persistent pain or pressure in the chest, pale, gray, or blue-colored skin, lips, or nail beds.^27 28 29^ Each participant is then prompted to collect a weekly nasal swab. As part of the weekly Symptom & Swabbing Survey, participants are asked whether they were tested for SARS-CoV-2 outside of the study and if they had a known exposure to COVID-19. Additionally, participants can report new symptoms when they occur outside their weekly symptom and swabbing survey reporting day.

### Weekly Illness Update and Recovery from Illness

Once an acute illness or positive test for SARS-CoV-2 is identified (Day 0), participants are asked to answer an Infection/Illness Onset Survey that captures specific symptoms, date of symptom onset, overall health status, and potential COVID-19 exposures. Following this survey, participants or their parent/guardian complete a post-infection/illness follow-up survey on Day 7 and Day 14 after they reported symptom onset. These surveys assess symptoms, recovery, overall health status, additional effects of their infection/illness (e.g., health care utilization and work and school absenteeism), and COVID-19 prevention behaviors. A survey at Day 30 following a positive SARS-CoV-2 test is completed to capture the continuation or resolution of symptoms, including illness/infection conditions, overall health status, and pediatric asthma/wheezing. Participants who test positive for influenza and RSV will also complete a 30-day post illness survey.

Additionally, positive SARS-CoV-2 study results trigger the serial collection of nasal swabs every 3 days for 21 days for both asymptomatic and symptomatic participants. If a participant reports a positive test for SARS-CoV-2 outside the study, they will be prompted to collect a study swab within 72 hours. If the study swab is positive for SARS-CoV-2, serial swabbing will be triggered.

### Monthly Survey and Semi-annual Survey

Following enrollment, updated job information, absenteeism, vaccination status, respiratory virus testing outside of the study, and overall health status are collected on a monthly basis. The monthly survey design involves rotating questions which may change during the study but could assess the receipt of influenza or other routine immunizations, participation in extracurricular activities, social and emotional burdens related to the pandemic, mitigation measures, and knowledge, attitudes, and perceptions about COVID-19 vaccines and therapeutics. The semi-annual survey will include additional post-COVID-19 conditions and lifestyle questions (Supplemental Table 3).

### Immunization Information Systems

Sites will use data from state immunization information registries to obtain vaccination information for participants, including the Washington State Immunization Information System (WA IIS) and Oregon Health Authority ALERT Immunization Information System (ALERT IIS). For participants enrolled in the KPNW health plan, vaccination history will also be obtained from medical records. Participants are consented prior to accessing these databases.

### Use of Electronic Medical Record Data

Medical records data from participants regarding COVID-19 hospitalizations, clinical respiratory virus testing, respiratory illness visits, and associated diagnostic tests and treatments will be used to supplement participant self-reported history of illness. Participants are consented at enrollment for medical release of information.

### Respiratory Specimens

#### Materials

Swab kits include instructions, a sealed nasal swab, a dry tube with a barcode sticker linked to the individual participant, a biohazard bag, and a pre-addressed return polymailer bag (International Air Transport Association-compliant Category B; Figure 3A, 3B, 3C). All specimen collection materials are FDA-approved or authorized for SARS-CoV-2 RT-PCR testing. Direct shipments of kits are sent on a regular basis to ensure a continuous supply of swabs available to participants. For households with multiple participants, the swab kit contains swabs for all household members. For these households, the collection kits for each household member are divided into separate boxes with labels used to designate each household member. At enrollment, each participant is asked to choose an animal, which is printed on a sticker that is used to label their swab kits and serve as a personal identifier throughout the study (Figure 3D). In addition, each swab has a barcode which must be entered into the online Symptom and Swabbing survey to activate the swab and ensure correct processing and linkage of results to an individual participant.

**Figure 3.**
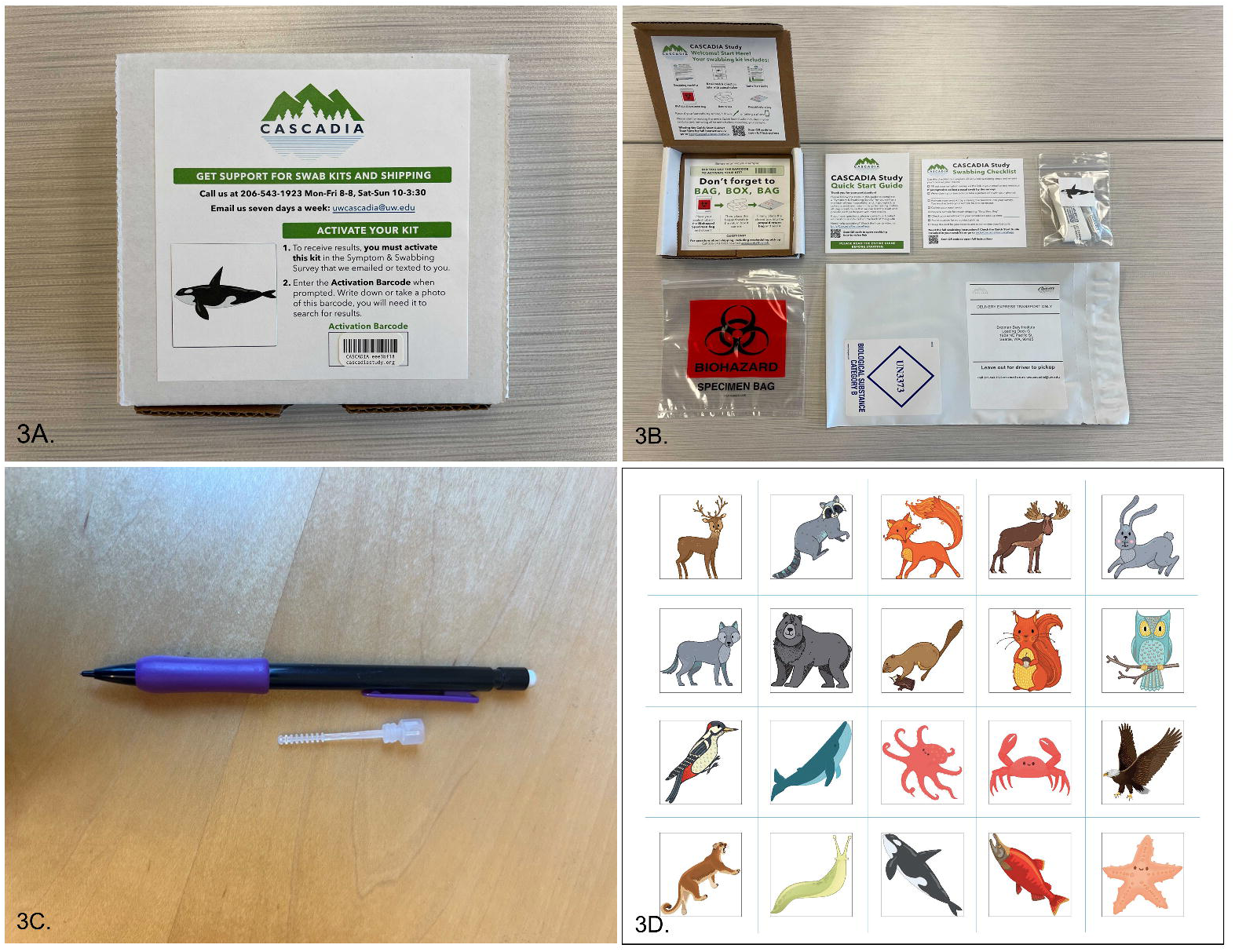
3A. Participant swab kit, 3B. Participant swab kit contents, 3C. TINY RHINOstic swab, 3D. Participant Stickers

#### Collection and Processing

TINY RHINOstic (anterior nares) (Rhinostics, Boston, MA) nasal swabs are being used for nasal specimen collection for all participants. For children aged 6 months to 12 years, a parent/guardian collects the nasal swab; swabs can be collected by a parent/guardian or self-collected under supervision of their parent/guardian for children aged 13 to 17.

All nasal swabs are dry and transported via courier service at ambient temperatures from participant households to the UW laboratory for processing. Specimens from the Portland, OR and Vancouver, WA area are transported to Seattle twice daily by courier. The limit of detection at 2-4 molecules/μL, includes 100% sensitivity, and 99.4% specificity for SARS-CoV-2.^30^ Swabs are processed at the lab within 48 hours of receipt. A participant communications team is available by phone seven days a week to assist participants.

#### Laboratory Testing

Nasal swab specimens are tested using an RT-PCR assay for SARS-CoV-2. All swabs are stored dry without media and eluted with 300μL Tris-EDTA for RHINOsticTM. 50μL of eluate is treated with proteinase K and heat for direct RT-qPCR (Swab-Express RT-qPCR).^31^ The RT-qPCR assay utilizes probe sets for SARS-CoV-2 Orf1b and S gene (Life Technologies) against ancestral virus and are multiplexed with probe against RNaseP and set on a QuantStudio 6 instrument (Applied Biosystems).^31^ The laboratory is clinically certified to test for SARS-CoV-2 by the Washington State Department of Health (WA DOH). As funding permits, specimens will undergo multiplex RT-PCR testing for SARS-CoV-2, influenza A/B and RSV as well as other respiratory pathogens.

Viral genome sequencing will be attempted on all SARS-CoV-2-positive samples with a Ct value of 35 or less using a hybrid capture method or a COVID-seq kit (Illumina). Raw sequencing reads will be processed using the Seattle Flu Assembly Pipeline (publicly available on GitHub).^32^ Sequences will be aligned, and phylogenetic trees constructed using the Nextstrain augur software. All assembled genomes will be made publicly available in the GISAID database and categorized according to lineage.

#### Return of Results

The UW laboratory will provide RT-PCR results for SARS-CoV-2. Testing results will be loaded into a REDCap module and linked to participant records that correspond to the barcoded specimen kits. In parallel, the testing results will be loaded into an online return of results portal. If a participant’s specimen tests positive for SARS-CoV-2, participants and or parents/guardians of participants will be contacted via text, email and/or phone for guidance on further study procedures. Positive SARS-CoV-2 results will be reported to local and state public health authorities to be compliant with notifiable disease reporting guidelines.

### Serum and Blood Specimens

#### Collection and Processing

All participants are asked to provide blood samples prior to enrollment and annually.

Participants who consent for sub-studies may undergo additional blood draws and mucosal sampling every six months and following immunizations and/or illness.

Blood volumes collected differ by participant age. We collect approximately 5 mL from children <13 years of age, and 20 mL from participants 13 years of age and older. For participants unable to provide a venous blood draw, the use of a Tasso (Tasso, Inc.) home blood collection device is under consideration.^33^

All samples are logged into REDCap using a blood sample collection form. Serum specimens are divided into aliquots labeled with the participant ID and an aliquot ID unique to each tube. At KPNW serum samples are temporarily stored at 4°C for up to 72 hours and subsequently stored in a -20°C or colder freezer prior to being shipped to UW laboratory. At UW or SCH, specimens are kept in a 4°C refrigerator for up to 96 hours at the clinical collection site and subsequently transported to the UW laboratory on wet ice by a courier service or by study staff. These samples are then aliquoted and stored at –20°C or colder in a temperature-monitored freezer until serologic testing is performed. After testing, residual specimens are stored at - 20°C or colder for long-term storage, and possible additional future testing in individuals in whom consent was obtained for additional testing. Aliquots may be sent to the CDC or to a CDC-designated laboratory for additional immunologic testing or long-term storage.

#### Laboratory Testing

Serum specimens will be tested at UW for antibodies against SARS-CoV-2 antigens, including nucleocapsid (N), spike (S), receptor binding domain (RBD), and potentially others.

Quantitative ELISA will be performed to test for IgG antibodies (Mesoscale diagnostics). A WHO standard will be included as part of these assays for standardization against an international standard.

Additional assays that may be performed in a subset of samples include influenza hemagglutinin inhibition assays, RSV neutralization assays and ELISAs, cytokine assays, flow cytometry and cytokine multiplex assays for characterization of the B and T-cell repertoire, as well as SARS-CoV-2 pseudovirus neutralization assays for VOCs and wild-type neutralization assays.

#### Participant Retention

Participants are allowed to pause weekly study activities for reasons such as family event or vacations, by notifying study staff. Text messages, emails, and phone calls that encourage participation are implemented at determined time intervals if participants do not complete weekly swabbing and symptom surveys for more than 2 consecutive weeks without notifying study staff. After five weeks of non-compliance with weekly swabbing and subsequent lack of response to study retention outreach, participants will be withdrawn from the study. Participants may be reenrolled if a future response is received.

#### Patient and Public involvement

Participants can provide feedback throughout the study. The study team reviews this feedback and study changes are made when possible. Study results are currently planned to be shared via interim newsletters and publications.

#### Remuneration

Participants are provided gift cards and compensated for completion of different study activities. They are compensated on a monthly basis, based on the completion of weekly swab collection and surveys. Participants receive further compensation for completing illness swab series and for visits for blood collection.

### Data Analysis

#### Vaccine Effectiveness

For the VE objectives, Cox regression models will be used to estimate the hazard ratio associated with SARS-CoV-2 vaccination, controlling for important demographic factors and potential confounders. COVID-19 VE will be calculated as:

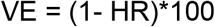

Cox regression models are well suited to account for censored person-time (as subjects disenroll) and time-varying exposures (as subjects become vaccinated or infected). Household-level clustering will be accounted for if more than one participant from the same household is enrolled in the study. VE will be estimated separately for the effectiveness in preventing symptomatic SARS-CoV-2 infection (defined by presence of symptoms and detection of SARS-CoV-2 by RT-PCR) and asymptomatic SARS-CoV-2 infection. VE will also be estimated for preventing medically attended SARS-CoV-2 infection; however, it is anticipated that power will not be adequate to calculate VE to prevent severe infection resulting in hospitalization, ICU admission, or death.

The effect of the following covariates on VE estimates will be considered for all outcomes: socio-demographic characteristics (e.g., age, sex, race, and ethnicity), exposures (school, day care, extracurriculars, social), study site, percent of other household members vaccinated, and community-level circulation of the virus. Stratified analyses will be conducted to estimate VE by age group, prior infection at enrollment (using seropositivity and self-reported infections), vaccine product, vaccine doses (primary series and boosters), and other factors, including time since last vaccination and prior infection. We acknowledge that this study is underpowered for pediatric VE and we will be exploring combining data with other CDC VE studies.

#### Incidence Objectives

The crude incidence rate will be calculated as the ratio of the number of SARS-CoV-2 infections divided by the total person-time contributed by all study participants during the follow-up period. Participants’ person-time will be censored at the time of first infection during the follow-up period or loss from cohort. The 95% confidence interval will be estimated assuming a Poisson distribution. Incidence will be presented overall, and separately, for symptomatic and asymptomatic laboratory confirmed SARS-CoV-2. Incidence will also be stratified by vaccination status, age, sex, race and ethnicity, history of prior infection, and other important risk factors. Funding permitted; the crude incidence rate will be calculated for other respiratory viruses.

#### Ethics and dissemination

All study materials including the protocol, informed consent forms, participant communications and recruitment materials, data collection instruments and other documents associated with the protocol were approved prior to study implementation. The KPNW IRB serves as the IRB of record for the study, with UW, SCH, and CDC ceding to the KPNW IRB

#### Assessment of harm and adverse events

Breaches of confidentiality, protocol deviations and adverse events are tracked by the study team. Events determined by the principal investigators to meet reporting criteria are submitted to the IRB.

## RESULTS

The study enrollment period began in June 2022 and enrollment is on-going. As of December 8, 2022, we have enrolled 2,047 persons from 788 households. Of enrolled participants, 59% of participants identify as female and 7% as Hispanic. Of the participants, 73% identify as White, 7% Asian, 2% Black/African American, 0.3% American Indian/Alaska Native, 0.1% Hawaiian/Pacific Islander, 9% multiple races, 1% other and 1% unknown. The majority of enrolled participants are adults 18-49 years (52%) followed by 5-11 years (26%), 12-17 years (14%), 2-4 years (6%), and 6 mos-1 year (2%).

## DISCUSSION

The CASCADIA study is enrolling participants in order to estimate the incidence of asymptomatic and symptomatic SARS-CoV-2 infection and real-world COVID-19 VE in children and adults. Additionally, our study allows for the evaluation of humoral immune response following vaccination and infection, estimation of household transmission of SARS-CoV-2 in children and adults, and monitoring of the evolution of the SARS-CoV-2 virus by genomic sequencing.

This study design has several strengths. The CASCADIA study uses a remote web-based survey model which allows for access to a larger population and captures sociodemographic, clinical and behavioral characteristics of persons before, during, and after a respiratory illness from SARS-CoV-2. The surveys are designed to collect comprehensive data but are not time consuming for participants. The four-year study design and collection of weekly respiratory samples will allow us a standardized methodology for capturing our primary outcome of VE, including both symptomatic and asymptomatic infection. This study allows for the collection of data regarding community-based transmission among children and adults in persons who might not have otherwise sought medical care or testing. Weekly nasal swab collection captures pre-symptomatic, symptomatic, and asymptomatic SARS-CoV-2 infection. Additionally, the inclusion of blood specimens will allow us to assess the kinetics and durability of immune response following infection and vaccination. By using multiplex molecular testing, immunity to variants of SARS-CoV-2 will be assessed. This assay was used previously to recognize community transmission in the beginning of the SARS-CoV-2 pandemic in the United States.^34 35^ Genomic sequencing of SARS-CoV-2 will provide information about circulating variants strains, intra-patient virus evolution, and changes in household or community transmission patterns. The rotating design of the monthly surveys allows for flexibility in data collection which may change as the study progresses.

In studies evaluating vaccine effectiveness in the pediatric population, medical care and symptomatic infection are typical endpoints.^36 37 38 39 40^ The CASCADIA study will collect longitudinal information in a pediatric and adult cohort using a weekly swabbing study design to evaluate the clinical spectrum SARS-CoV-2 infection, spanning asymptomatic viral shedding to severe disease. The prospective design and active surveillance within the study allows for the identification of risk factors leading to symptomatic and asymptomatic infection.

This study has limitations. Given the study design, a selection bias of pro-medical and vaccinated persons will likely occur. Enrollment of persons with distrust of the medical community may be difficult and under-enrollment of unvaccinated persons is expected. The web-based survey design may introduce selection bias and limit generalizability. Recruitment efforts will be adapted throughout the enrollment period to ensure a diversity of study participants. Given the longitudinal nature of the study, retention and consistency will need to be monitored and carefully maintained. Annual blood collection may be a limitation for child participants and if a hinderance to enrollment, this mandatory requirement may require re-consideration. Additional blood collection methods such as a Tasso (Tasso, Inc.) home blood collection device is being explored. ^33^

In conclusion, this study design allows for large population surveillance of SARS-CoV-2 and other respiratory viruses with a predominantly remote, web-based structure to inform on real world vaccine effectiveness in children and adults.

## Data Availability

All data produced in the present study are available upon reasonable request to the authors

https://www.cascadiastudy.org

## Acknowledgements

Deralyn Almaguer, Britt Ash, Kristi Bays, Tara Beatty, Trevor Bedford, Kristin Bialobok, Allison Bianchi, Cathleen Bourdoin, Stacy Bunnell, Joseph Cerizo, Evelin Coto, Phil Crawford, Lantoria Davis, Lisa Fox, Kenni Graham, Tarika Holness, Madison Hollcroft, Matt Hornbrook, Keelee Kloer, Dorothy Kurdyla, Max Lin, Natalie Lo, Kyle Lutein, Richard Martin, Melissa P. MacMillan, Ariana Magedson, Denise McCulloch, John Ogden, Aaron Piepert, Joanne Price, Angela Reyes-Ochoa, Jennifer Rivelli, Sperry Robinson, Katrina Schell, Emily Schield, Jay Shendure, Anna Shivinsky, Valencia Smith, Jeremy Stone, Alexandra Varga, Mica Werner.

## Contributorship Statement

All authors contributed to the study protocol and design. All authors contributed to manuscript revisions and approved the final version of the manuscript.

