## Supplemental Tables for "CASCADIA: A prospective community-based study protocol for assessing SARS-CoV-2 vaccine effectiveness in children and adults utilizing a remote nasal swab collection and web-based survey design"

Table 1. Exploratory Objectives

Exploratory Study Objectives

1. Examine the incidence and outcomes of other respiratory virus infections, including respiratory syncytial virus (RSV) and influenza virus infection.
2. Estimate the incidence of co-infection with SARS-CoV-2 virus with other respiratory viruses in children and adults and identify associated risk factors.
3. Estimate VE of influenza vaccines in preventing influenza virus infection and influenza illness.
4. Estimate effectiveness of respiratory virus pharmaceutical preventative interventions that are available during the timeline of the study.

Exploratory Immunologic Objectives

1. Evaluate concordance between serologic (neutralizing antibody, anti-N, anti-S, anti-RBD ELISA) and molecular tests (SARS-CoV-2 RT-PCR) for diagnosis of symptomatic and asymptomatic infection in children and adults.
2. Evaluate concordance between different serologic assays (neutralizing antibody, anti-N, anti-S, anti-RBD ELISA) for diagnosis of symptomatic and asymptomatic infection in children and adults.
3. Estimate influenza VE in children and adults against serologic and virologic-confirmed infection.
4. Estimate the incidence of serologic influenza infection in children and adults and identify associated risk factors for seroconversion.
5. Evaluate the effect of baseline influenza haemagglutinin-inhibiting antibodies (HAI) and other antibody titers on risk of infection with influenza, and evaluate factors including symptom status, age, and virologic factors (strains, viral load).
6. Evaluate the kinetics and half-life of influenza HAI and other antibody in children and adults, and identify factors associated with increased half-life.
7. Evaluate the effect of annual doses of influenza vaccine on HAI and other antibody titers.
8. Estimate the incidence of serologic infection with respiratory pathogens in children and adults and identify associated risk factors for seroconversion.
9. Evaluate the effect of baseline antibody titers on risk of infection and evaluate factors including symptom status, age, and virologic factors (strains, viral load) for multiple respiratory pathogens.
10. Evaluate the kinetics and half-life of antibody in children and adults and identify factors associated with increased half-life for multiple respiratory pathogens.
11. Explore the role of mucosal immune responses in response to infection for multiple respiratory pathogens.
12. Evaluate the role of specific Th1 and Th2 cytokines in response to infection for multiple respiratory pathogens.
13. Evaluate the cross-protective effect of the immune response to respiratory pathogens to subsequent infections (e.g., SARS-CoV-2 immunity against subsequent human coronavirus infections).

| Table 2. Enrollment Survey Data |
|---|
| Socio-demographic characteristics<br>sex-assigned at birth<br>gender<br>race<br>ethnicity<br>health insurance coverage<br>employment information<br>income bracket |
| Residence characteristics<br>Type of home<br>Household composition |
| Household Data<br>Age of all household members<br>COVID-19 and influenza vaccination status of each household member |
| Health history overall<br>Overall health status<br>Self-reported medical conditions<br>Vaccination history (SARS-CoV-2 and other selected vaccines)<br>Smoking history |
| In-person school, childcare, and work attendance |
| Participation in extracurricular, work-related, and social activities |
| Mask-wearing and other mitigation measures for children over 2 years of age |
| Previous history of positive SARS-CoV-2 testing |
| Vaccine knowledge, attitudes, and perceptions |

Table 3 Scheduled Survey Contents for All participants

| | Scheduled Surveys | | | | |
|---|---|---|---|---|---|
| **Variable** | **Screener** | **Enrollment** | **Weekly Symptom & Swabbing Survey** | **Monthly Survey** | **Semi-Annual Survey** |
| Contact Information | X | | | | |
| Demographics | X | X | | | |
| Income and Education | | X | | | |
| Household information | | X | | X | |
| Recruitment Source | X | | | | |
| Occupation related questions[1] | | X | | X | X |
| School related questions | | X | | | X |
| Sick Leave Policy | | X | | X | X |
| Healthcare Access | | X | | X | |
| Height/weight | | X | | | |
| Comorbidities | | X | | | X |
| General Health | | X | | X | X |
| COVID-like symptoms | | X | X | | X |
| Pediatric wheezing assessment[2] | | X | | | X |
| Smoking assessment[3] | | X | | | X |
| Influenza positive testing history | | | X | | |
| COVID-19 positive testing history | | X | X | | |
| COVID-19 hospitalization history | | X | | | |
| COVID-19 treatment history | | X | | | |
| COVID-19 Vaccines | X[4] | X | | X | |
| Vaccine perceptions | | X | | X* | |
| Influenza vaccines | | X | | X | |
| Social Activities | | X | | X* | |
| Masking practices | | X | | X* | |
| Absenteeism | | | | X | |
| New Medical Diagnoses | | | | X | |
| Enrollment clinical trial/investigation treatment COVID-19 | | | | X* | |
| COVID-19 preventative medications | | | | X* | |
| COVID-19 perceptions | | | | X* | |
| Childcare | | | | | X |
| RSV knowledge, attitudes and practices questions | | | | X | |

1:$\geq$ 18 years, 2: $\leq$13 years old, 3: $\geq$12 years old, 4: only total number of vaccines, * rotating questions

Table 4. Study Activities

| | Study activities | | | | | | | |
|---|---|---|---|---|---|---|---|---|
| **Study Activity** | **At Enrollment** | **Weekly** | **Monthly** | **As needed if the following events happen:** | | | **Every 6 Months** | **Once a year** |
| | | | | **If COVID-19 symptoms** | **If COVID-19 and/or Flu Vaccine** | **If tests positive for COVID-19, Flu or other virus** | | |
| Review, sign, and date consent document | ✓ | | | | | | | |
| Enrollment Survey | ✓ | | | | | | | |
| Monthly Update Survey | | | ✓ | | | | | |
| Symptom and Swabbing Survey | | ✓ | | ✓ | | | | |
| Illness Surveys | | | | ✓ | | ✓ | | |
| 6 Month Update Survey | | | | | | | ✓ | |
| Nasal Swab | | ✓ | | ✓ (if more than 48 hours after weekly swab) | | ✓ (every 3 days for 3 weeks)[1] | | |
| Blood draw | 🩸 | | | | | | | 🩸 |

| | Study activities | | | | | | | |
|---|---|---|---|---|---|---|---|---|
| **Study Activity** | **At Enrollment** | **Weekly** | **Monthly** | **As needed if the following events happen:** | | | **Every 6 Months** | **Once a year** |
| | | | | **If COVID-19 symptoms** | **If COVID-19 and/or Flu Vaccine** | **If tests positive for COVID-19, Flu or other virus** | | |
| Optional blood draw for Sub-study | 🩸 | | | | | | | 🩸 |
| Optional blood draw for Sub-study | | | | | 🩸* | 🩸* | 🩸 | |
| Optional saliva and additional nasal swab for optional Sub-study | ✓ | | | | ✓ | ✓ | ✓ | |

[1]SARS-CoV-2 positive only, * up to 30 days post-vaccine or infection. Planned laboratory studies from blood samples include antibody and neutralization assays, and B and T cell assays from collected PBMCs. Planned laboratory studies from nasal swabs include SARS-CoV-2 RT-PCR and genomic sequencing from positive SARS-CoV-2 samples. Secondary and exploratory studies include RT-PCR for Influenza and RSV, antibody and neutralization assays for other respiratory viruses, OpenArray™, and hemagglutination inhibition assay. From the optional mucosal sub-study samples, IgG, IgA, and neutralization assays studies are planned.

Optional Sub-studies

Three optional sub-studies are planned for CASCADIA. An additional volume of blood will be requested from a subset of participants, both adult and pediatric, on a voluntary basis, for collection. Sub-studies include: 1) additional blood collection for antibody testing at enrollment and annually 2) sera and peripheral blood mononuclear cells (PBMCs) and whole blood for additional profiling, including T and B-cell profiling assays at enrollment, post vaccine or infection, or every 6 months 3) mucosal sampling (nasal and saliva) for mucosal serologic immune assays.

PBMC isolation will be performed using Ficoll-Hypaque density gradient centrifugation and resuspended in freezing medium containing 90% fetal bovine serum, and 10% DMSO. Cells will then be transferred to cryogenic vials and placed in cryofreezing containers and stored at –80 $^{o}$C for 24-48 hours, and subsequently transferred to liquid nitrogen freezers and stored in the vapor phase at –132 $^{o}$C or colder until use.

In a subset of individuals, both adult and pediatric, we may additionally collect nasal or salivary secretions for additional assays to evaluate the role of mucosal immunity in protection from and in response to infection and vaccination. Participants in this optional study will collect nasal swab and saliva specimens at enrollment, post vaccine or infection, or every 6 months.
